# Temporal Response and Survival Dynamics After Durvalumab in Stage III NSCLC: A Response-Augmented Continuous-Time Markov Framework

**DOI:** 10.1101/2025.10.01.25337053

**Authors:** A.J. Wals Zurita, M.A. González Ruíz, D. Muñoz Carmona, J. Pachón Ibáñez, G. Campos Rivera, C. Míguez Sánchez

## Abstract

**Background:** Progression-free survival combines clinically different experiences, including objective tumor response and disease control without response. We developed an aggregate-data model to estimate time spent in these states after durvalumab consolidation for unresectable stage III non–small-cell lung cancer.

**Methods:** A piecewise continuous-time illness–death model was constructed from paired overall-survival (OS) and progression-free-survival (PFS) landmarks from the durvalumab arm of PACIFIC at 12, 24, and 36 months. Outcomes at 48 and 60 months were reserved for temporal validation. Progression-free occupancy was partitioned into ongoing objective response and disease control without ongoing response by convolving an explicit response-detection distribution with the published duration-of-response survival function. The primary analysis assumed responses were detected from month 2, reached 28.4% by month 12, and reached the mature 29.8% objective response rate by month 24. Earliest-detection, assessment-scheduled, and full-cohort-denominator scenarios assessed structural uncertainty. A partial landmark-uncertainty analysis sampled published OS/PFS intervals and response counts while holding duration-of-response landmarks fixed. Restricted mean state times were integrated to 60 months.

**Results:** Predicted versus observed 60-month OS was 42.5% versus 42.9%, and PFS was 30.9% versus 33.1%. Temporal-validation root mean squared error at 48–60 months was 0.60 percentage points for OS and 1.56 points for PFS. At 60 months, estimated occupancy was 15.6% in ongoing objective response, 15.3% in progression-free control without response, 11.6% in progressed disease, and 57.5% in death. Restricted mean times over 60 months were 10.92, 17.69, 10.71, and 20.69 months, respectively; partial plausibility ranges were 9.87–12.42, 14.75–20.03, 8.31–13.66, and 18.61–22.55 months. Response time was 10.91 months with assessment-scheduled detection, 11.78 months with earliest detection, and 10.16 months when the responder count was divided by the full randomized cohort.

**Conclusions:** Published response and survival summaries support estimation of time in ongoing objective response as a clinically interpretable subdivision of PFS. They do not support a fully parameterized model of transitions among complete response, partial response, stable disease, post-response relapse, progression, and death. Reporting the estimable four-state occupancy together with structural scenarios and partial plausibility ranges preserves clinical detail without implying unsupported precision. The trajectories may provide exploratory aggregate inputs for planning only when combined with local activity and resource-use data.

**Highlights:**

- Published survival and response data were combined in one state-occupancy framework.
- The model estimates time in ongoing objective response rather than only progression-free survival.
- Mean 60-month occupancy was 10.92 months in response and 17.69 months in nonresponse disease control.
- Complete- and partial-response times were not separately identifiable from aggregate data.
- Time-varying occupancy can provide exploratory cohort-level planning inputs when combined with local resource data.
- Response-onset scenarios and executable code make the structural assumptions explicit.

## 1. Introduction

Durvalumab consolidation after platinum-based concurrent chemoradiotherapy improves progression-free survival (PFS) and overall survival (OS) in patients with unresectable stage III non–small-cell lung cancer (NSCLC) without progression [1–4]. Five-year follow-up of the PACIFIC trial showed that 42.9% of durvalumab-treated patients were alive and 33.1% remained progression-free [3]. These long-term proportions are important, but survival curves alone do not describe how progression-free time is experienced clinically. Patient-reported symptoms, functioning, and global health status or quality of life remained stable, without clinically meaningful differences between durvalumab and placebo [5].

Radiologic response categories provide a more granular description. Complete response (CR), partial response (PR), stable disease (SD), and progressive disease (PD) are defined from longitudinal tumor measurements and confirmation rules [6]. In PACIFIC, tumor assessments were scheduled every 8 weeks during the first year and every 12 weeks thereafter [1]. Among 443 durvalumab-treated patients with measurable disease, the initial analysis reported 6 CRs and 120 PRs, for an objective response rate (ORR) of 28.4% [1]. With extended follow-up, 132 responses were reported (ORR 29.8%), and 51.1% of responders remained in response at 60 months after response onset [3].

These publications contain enough information to quantify ongoing objective-response occupancy, but not every transition that a detailed clinical diagram might contain. In particular, aggregate reports do not provide patient-level response-onset times, state-specific durations for CR and PR, movements between CR and PR, or the timing of relapse after CR. Treating each of those pathways as separately estimated would therefore introduce parameters unsupported by the data.

We developed a response-augmented continuous-time Markov framework that uses all identifiable information while separating observations from assumptions. The objectives were to: (1) reproduce OS and PFS using a valid piecewise illness–death model; (2) partition PFS into ongoing objective response and disease control without ongoing response; (3) estimate restricted mean time in response, nonresponse disease control, progressed disease, and death; (4) quantify sensitivity to the unreported timing and denominator of response; and (5) examine how time-varying cohort occupancy could support operational planning without being interpreted as individual prediction.

## 2. Materials and Methods

### 2.1 Study design and data sources

This was a methodological study using published aggregate data from the durvalumab arm of PACIFIC. Paired OS and PFS estimates at 12, 24, and 36 months were used for parameter derivation [3]. The 48- and 60-month estimates were not used in construction and served as temporal validation. PACIFIC-R estimates at 24 and 36 months were retained as a contextual external comparison [7].

Response information came from the initial PACIFIC report and the five-year online appendix [1,3]. The initial report supplied assessment frequency, the number of evaluable patients, and the early distribution of best overall response. The mature report supplied ORR and the proportions of responders remaining in response at 12, 24, 36, 48, and 60 months. Values and analytical roles were specified before model execution (Supplementary Table S1).

### 2.2 Survival component

The survival component had three states: progression-free (PF), progressed disease (P), and death (D), corresponding to a progressive illness–death structure [8]. All patients entered PF at randomization. Within interval i, the generator was

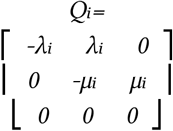

where λ_i_ is the monthly PF-to-P intensity and μ_i_ is the monthly P-to-D intensity. Transition probabilities were obtained from P_i_(t)=exp(Q_i_ t). PFS equaled PF occupancy, and OS equaled PF plus P occupancy.

Knots were placed at 12 and 24 months. Within each interval, λ_i_ was obtained from consecutive PFS landmarks. Conditional on λ_i_ and the incoming state vector, μ_i_ was solved by bisection so that the paired OS landmark was reproduced. Although state propagation has a closed-form expression for fixed transition rates, inversion with respect to μ_i_ is transcendental; bisection provides a stable solution to this monotone one-dimensional calibration problem. The rates estimated from months 24–36 were held constant after month 36. The 48- and 60-month landmarks were deliberately reserved for temporal validation; adding a further post-36-month interval would have consumed validation information. This is a marginal bookkeeping model: because the published aggregate curves do not distinguish death before recorded progression, PFS failures are represented as entering P before death. This restriction avoids introducing transition parameters that cannot be uniquely identified from the available calibration targets [9].

### 2.3 Response augmentation

Objective response was defined as CR or PR. Let F_r_(u) denote the assumed cumulative distribution of detected response onset from randomization and S_DOR_(v) the published conditional probability that a response remains ongoing v months after onset. Ongoing response occupancy was

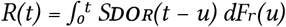

The conditional duration-of-response curve was interpolated with piecewise-constant hazards between the published 12-month landmarks. The corresponding monthly response-exit rates were 0.01746, 0.01227, 0.01467, 0.00851, and 0.00304 for durations 0–12, 12–24, 24–36, 36–48, and 48–60 months. An exit from ongoing response was not interpreted as a specific biological transition because the published data do not distinguish loss of response, progression, death, or assessment-related censoring at the individual level.

The primary response-onset scenario reflected the scheduled first assessment at approximately 2 months. Cumulative detected response increased linearly from 0 at month 2 to the early ORR of 28.4% at month 12, then to the mature ORR of 29.8% at month 24, and remained constant thereafter. Month 12 and month 24 are explicit modeling anchors rather than published response-onset quantiles. Linear interpolation was chosen as a parsimonious smoothed bridge between anchors, not as a literal representation of responses occurring continuously between discrete assessments.

The four mutually exclusive cohort occupancies were:

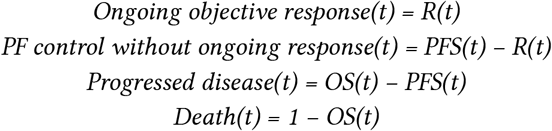

“PF control without ongoing response” includes SD, patients not yet classified as responders, and patients whose objective response ended without a separately observable progression transition. It must not be interpreted as a pure SD state.

Figure 1 summarizes the aggregate survival transitions and the descriptive partition of PFS. The dashed PFS boundary emphasizes that its two internal categories are occupancy components rather than separately fitted Markov transitions.

**Figure 1.**
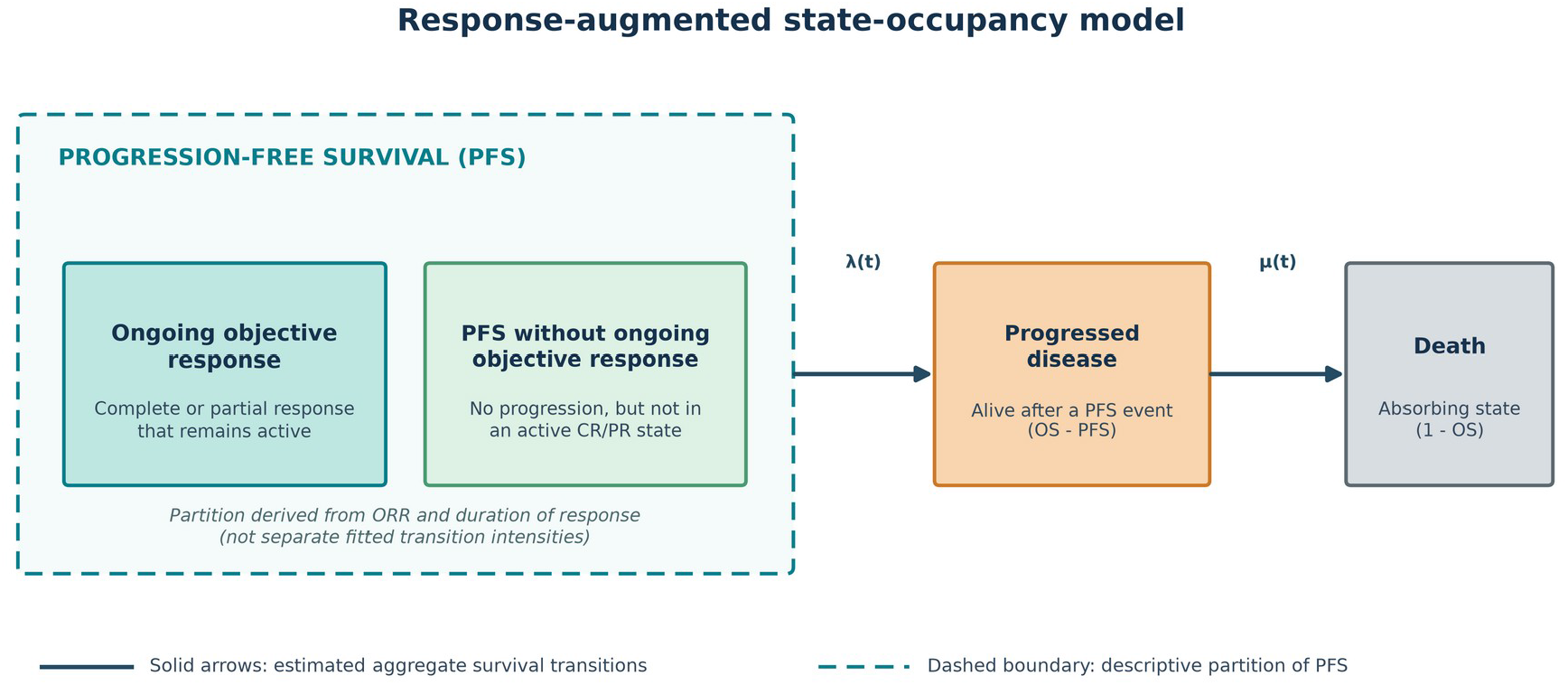
Structure of the response-augmented state-occupancy model. The progression-free region is partitioned into ongoing objective response (active CR/PR) and PFS without ongoing objective response (alive without progression but not currently in active CR/PR). Progressed disease denotes patients alive after a PFS event; death is absorbing. Solid arrows are estimated aggregate survival transitions. The dashed boundary is an ORR/DOR-based partition of PFS, not separately fitted transition intensities.

### 2.4 Sensitivity analyses

Three structural alternatives were prespecified. In the earliest-detection scenario, all mature responses occurred at month 2. In the assessment-scheduled scenario, equal portions of the early 28.4% ORR were assigned at the 2-monthly assessments through month 12, and equal portions of the subsequent increase to 29.8% were assigned at 3-monthly assessments through month 24. In the denominator scenario, the 132 mature responses were divided by all 476 randomized durvalumab patients (27.7%) rather than the 443 patients with measurable disease.

The measurable-disease denominator was retained for the primary response estimand because it matches the population in which PACIFIC reported ORR. Dividing the response count by all randomized patients implicitly classifies patients without measurable disease as nonresponders; it was therefore retained as a conservative full-cohort sensitivity analysis rather than used as the primary analysis. CR and PR were not modeled or reported as separate state times because response-specific onset and duration distributions were unavailable.

### 2.5 Outcomes and verification

The primary outcome was restricted mean time in each state through 60 months, calculated by numerical integration of occupancy. State occupancy at prespecified landmarks and temporal-validation root mean squared error (RMSE) at months 48 and 60 were secondary outcomes. Automated tests required nonnegative occupancy, response occupancy no greater than PFS, exact summation of states to one, exact reproduction of the parameter-derivation landmarks by construction, reproduction of duration-of-response landmarks, and restricted state times summing to the analysis horizon.

Partial landmark uncertainty was explored with 3,000 accepted Monte Carlo draws using a fixed random seed. Published pointwise 95% intervals for the six OS/PFS parameter-derivation landmarks were approximated on the logit scale and sampled independently; draws were retained only if OS and PFS were nonincreasing and PFS did not exceed OS. Initial and mature ORRs were sampled from Jeffreys beta distributions based on the reported response counts, with mature ORR constrained not to be lower than initial ORR. Duration-of-response landmarks were held fixed because their joint uncertainty was not reported. The 2.5th and 97.5th percentiles are reported as partial plausibility ranges, not confidence intervals, because landmark covariance, uncertainty in duration of response, and uncertainty in the response-onset distribution could not be fully propagated.

All calculations were implemented in Python with NumPy and Matplotlib. Source data, code, tests, and machine-readable results accompany the study.

## 3. Results

### 3.1 Survival model

The monthly PF-to-P intensity declined from 0.04877 at 0–12 months to 0.01778 at 12–24 months and 0.01044 after month 24. Corresponding P-to-D intensities were 0.07859, 0.05788, and 0.04205 month^{-1} (Table 1). By construction, the interval-specific rates reproduced the six landmarks used for parameter derivation; this exact agreement is not a validation result.

**Table 1.** Monthly transition intensities in the survival component.

| Interval (months) | PF→progressed | Progressed→death |
| --- | --- | --- |
| 0–12 | 0.048766 | 0.078591 |
| 12–24 | 0.017776 | 0.057880 |
| ≥24 | 0.010443 | 0.042050 |

At 48 months, predicted OS was 48.9% versus 49.7% observed, and predicted PFS was 35.0% versus 35.0% observed. At 60 months, predicted OS was 42.5% versus 42.9% observed, and predicted PFS was 30.9% versus 33.1% observed. Temporal-validation RMSE was 0.60 percentage points for OS and 1.56 points for PFS (Figure 2).

**Figure 2.**
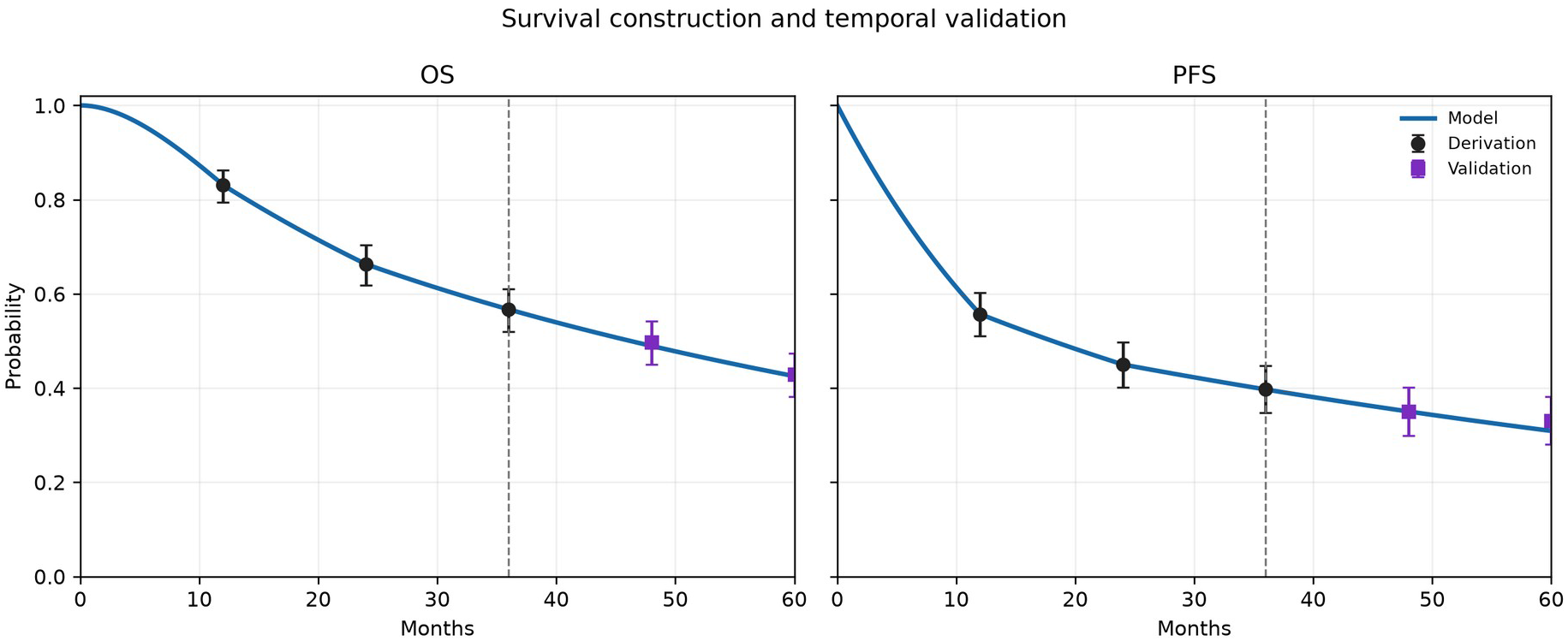
Survival construction and temporal validation. Circles denote parameter-derivation landmarks through 36 months; squares denote 48- and 60-month observations reserved for validation. Exact agreement with the circles is imposed by construction.

### 3.2 Response-augmented occupancy

Under the primary onset scenario, ongoing objective-response occupancy was 26.1% at month 12, 22.9% at month 24, 19.6% at month 36, 16.9% at month 48, and 15.6% at month 60 (Figure 3). At month 60, the remaining cohort occupancy was 15.3% in PF control without ongoing response, 11.6% in progressed disease, and 57.5% in death. Over the 60-month horizon, restricted mean time was 10.92 months in ongoing objective response, 17.69 months in PF control without ongoing response, 10.71 months in progressed disease, and 20.69 months in death (Table 2). Thus, restricted mean PFS time was 28.61 months, restricted mean post-progression survival was 10.71 months, and restricted mean OS time was 39.31 months.

**Table 2.** Restricted mean state time through 60 months.

| State | Point estimate, months | Partial plausibility range, months |
| --- | --- | --- |
| Ongoing objective response | 10.92 | 9.87–12.42 |
| PF control without ongoing response | 17.69 | 14.75–20.03 |
| Progressed disease | 10.71 | 8.31–13.66 |
| Death | 20.69 | 18.61–22.55 |

**Figure 3.**
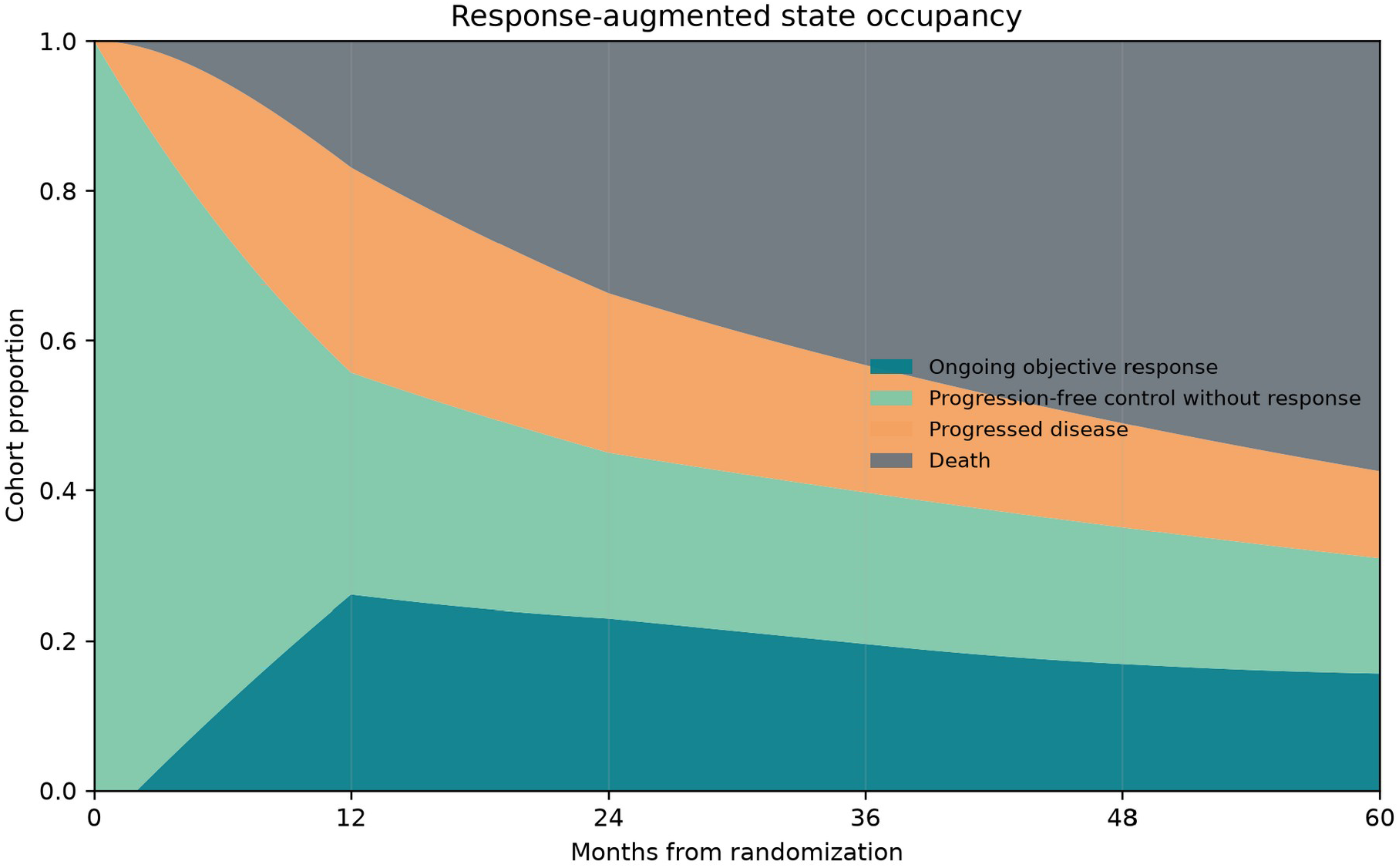
Response-augmented state occupancy. Ongoing objective response, progression-free control without response, progressed disease, and death sum to one at every time.

Values are rounded independently and may not sum to exactly 60.00 months. The ranges reflect sampled OS/PFS landmarks and response-count uncertainty with duration-of-response landmarks held fixed; they are not formal confidence intervals.

### 3.3 Structural sensitivity

Restricted mean objective-response time was 11.78 months when all responses were assigned to the earliest assessment and 10.91 months when detection was restricted to scheduled assessment times (Figure 4). Applying the mature responder count to the entire randomized durvalumab arm produced 10.16 months. The near agreement between the smoothed primary and assessment-scheduled scenarios indicates that discretizing response detection at visits had little effect on the 60-month restricted mean; denominator choice had a larger effect.

**Figure 4.**
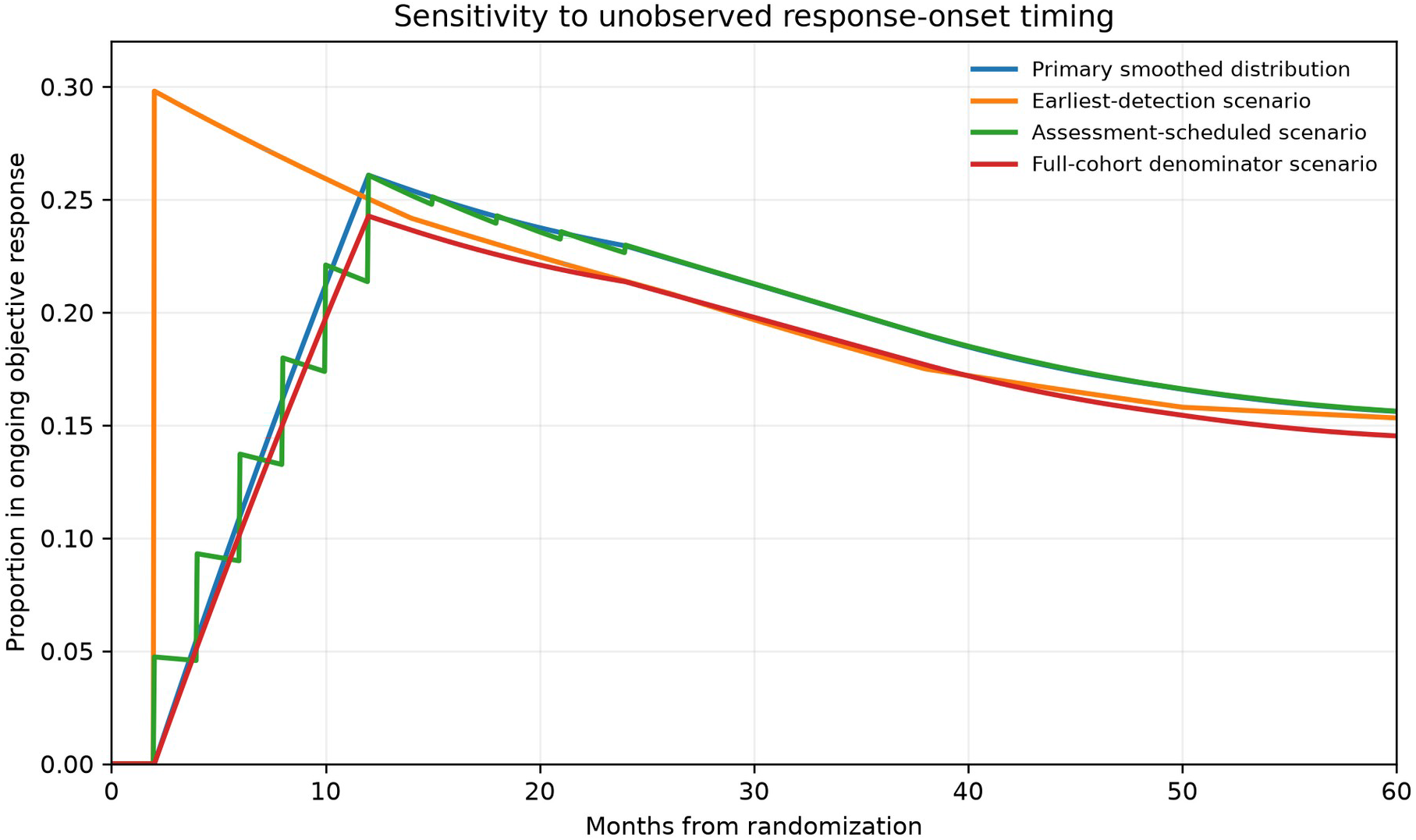
Sensitivity to response-onset timing and denominator. Ongoing objective-response occupancy under the primary smoothed, earliest-detection, assessment-scheduled, and full-cohort-denominator scenarios.

## 4. Discussion

This analysis translates published PACIFIC response and survival summaries into clinically interpretable time spent in four mutually exclusive states. During the first five years after randomization, the model estimated an average of 10.9 months in ongoing objective response and 17.7 months in progression-free control without ongoing response. Approximately half of the modeled PFS time was therefore spent in an active CR/PR category under the primary assumptions.

The time-indexed trajectories can also bridge clinical evidence and service logistics, but only at an exploratory cohort level. Progression-free occupancy may help frame demand for surveillance imaging and longitudinal oncology review, while modeled progression flow and post-progression occupancy may inform capacity for diagnostic work-up, multidisciplinary reassessment, salvage therapy, palliative radiotherapy, and supportive care. The residual progression-free state is heterogeneous and cannot be mapped one-to-one to a specific visit type or resource. Multiplying occupancy and transition flow by expected cohort arrivals could yield approximate workload profiles only after adding local referral volumes, assessment schedules, pathways, and resource intensity; the model itself does not estimate resource use. These occupancy profiles should not be interpreted as health-state utilities; in NSCLC, utility may vary with time to death and requires separate patient-reported data [10].

The distinction matters because PFS alone treats radiologic response and stable disease as equivalent. For patients and clinicians, time in documented tumor response may convey a different experience from nonprogressive disease without response. The proposed convolution uses the reported ORR to determine how many patients can enter response and the duration-of-response curve to determine how long those responses remain active. It thereby adds response information without pretending that a full multistate transition history was published.

The model also defines the boundary of what can be learned. Separate CR and PR states would require their onset times, duration distributions, and movements between categories. A post-CR relapse state would require the number and timing of such relapses. None is available by randomized arm in the aggregate reports. The observed CR/PR count ratio is therefore retained only as descriptive evidence and is not converted into separate state-time estimates.

Response-onset timing is the main unobserved component. PACIFIC reported assessment frequency and landmark ORRs, not a cumulative incidence curve of first response. The primary smoothed distribution is consequently an assumption. Its influence was made visible through earliest-detection and assessment-scheduled scenarios rather than hidden in fitted transition rates. The near equality of the primary and assessment-scheduled restricted means is reassuring for the 60-month summary, but it does not validate the assumed month-by-month response curve.

The survival component performed well in temporal validation at 48 and 60 months after parameter derivation through month 36. This supports its use as an accounting backbone over five years, although the PFS prediction at month 60 remained 2.2 percentage points below the observed estimate. This residual may reflect late changes in progression risk not captured by holding the 24–36-month rates constant after month 36. Piecewise hazards summarize changing cohort risk and should not be interpreted as fixed individual biological rates. Alternative flexible parametric and cure models may be preferable for lifetime extrapolation or economic evaluation [11–13].

Limitations include reliance on published aggregate rather than patient-level data; lack of covariance among survival and response landmarks; uncertain response-onset timing; use of response data from the measurable-disease subset alongside survival data from the randomized cohort; and the inability to assign exits from response to SD, progression, or death. The partial uncertainty analysis treats pointwise OS/PFS landmarks as independent after imposing basic logical constraints and holds duration-of-response landmarks fixed. Its ranges describe sensitivity to the sampled inputs rather than repeated-sampling confidence intervals and probably understate total uncertainty. No placebo-arm model was fitted, and no causal effect of durvalumab on state time is claimed. The reported state times are descriptive occupancies within the durvalumab arm and must not be interpreted as differences or gains relative to placebo.

The framework is portable to other settings in which OS, PFS, ORR, and duration of response are published. Its central requirement is that every modeled quantity be linked to an observable marginal endpoint or clearly labeled assumption. Patient-level event histories would allow extension to genuine CR, PR, SD, progression, and death transitions using multistate methods.

## 5. Conclusions

Aggregate PACIFIC data support a response-augmented four-state occupancy analysis comprising ongoing objective response, progression-free control without ongoing response, progressed disease, and death. The estimated mean time in ongoing objective response during the first five years was approximately 11 months, with a partial landmark-uncertainty plausibility range of 9.87–12.42 months. Separate complete-response, partial-response, stable-disease, and post-complete-response relapse pathways remain beyond what the published aggregate data can identify. This distinction preserves the clinical value of response states while avoiding unsupported transition estimates. The resulting temporal profiles may support exploratory cohort-level planning of surveillance and post-progression services only when combined with local activity and resource-use data.

## Data Availability

All data produced in this work are contained in the manuscript.

## Declarations

### Ethics approval

Not applicable. Only aggregate data from published reports were used.

### Consent for publication

Not applicable.

### Data and code availability

All transcribed landmarks, executable code, automated tests, machine-readable outputs, and figures are provided in the accompanying reproducibility package.

### Author contributions

Conceptualization: A.J.W.Z. and M.A.G.R.; methodology: A.J.W.Z. and C.M.S.; software and formal analysis: A.J.W.Z.; validation: J.P.I., D.M.C., and C.M.S.; investigation: A.J.W.Z., M.A.G.R., and G.C.R.; writing—original draft: A.J.W.Z., M.A.G.R., and G.C.R.; writing—editing: all authors; supervision: A.J.W.Z.

## References

1. Antonia SJ, Villegas A, Daniel D, et al. Durvalumab after chemoradiotherapy in stage III non-small-cell lung cancer. N Engl J Med. 2017;377:1919–1929. doi:10.1056/NEJMoa1709937.

2. Antonia SJ, Villegas A, Daniel D, et al. Overall survival with durvalumab after chemoradiotherapy in stage III NSCLC. N Engl J Med. 2018;379:2342–2350. doi:10.1056/NEJMoa1809697.

3. Spigel DR, Faivre-Finn C, Gray JE, et al. Five-year survival outcomes from the PACIFIC trial: durvalumab after chemoradiotherapy in stage III non-small-cell lung cancer. J Clin Oncol. 2022;40:1301–1311. doi:10.1200/JCO.21.01308.

4. Gray JE, Villegas A, Daniel D, et al. Three-year overall survival with durvalumab after chemoradiotherapy in stage III NSCLC: update from PACIFIC. J Thorac Oncol. 2020;15:288–293. doi:10.1016/j.jtho.2019.10.002.

5. Hui R, Özgüroğlu M, Villegas A, et al. Patient-reported outcomes with durvalumab after chemoradiotherapy in stage III, unresectable non-small-cell lung cancer (PACIFIC): a randomised, controlled, phase 3 study. Lancet Oncol. 2019;20:1670–1680. doi:10.1016/S1470-2045(19)30519-4.

6. Eisenhauer EA, Therasse P, Bogaerts J, et al. New response evaluation criteria in solid tumours: revised RECIST guideline (version 1.1). Eur J Cancer. 2009;45:228–247. doi:10.1016/j.ejca.2008.10.026.

7. Filippi AR, Bar J, Chouaid C, et al. Real-world outcomes with durvalumab after chemoradiotherapy in patients with unresectable stage III NSCLC: interim analysis of overall survival from PACIFIC-R. ESMO Open. 2024;9:103464. doi:10.1016/j.esmoop.2024.103464.

8. Meira-Machado L, Sestelo M. Estimation in the progressive illness-death model: a nonexhaustive review. Biom J. 2019;61:245–263. doi:10.1002/bimj.201700200.

9. Alarid-Escudero F, MacLehose RF, Peralta Y, Kuntz KM, Enns EA. Nonidentifiability in model calibration and implications for medical decision making. Med Decis Making. 2018;38:810–821. doi:10.1177/0272989X18792283.

10. Hatswell AJ, Chaudhary MA, Monnickendam G, et al. Modelling health state utilities as a transformation of time to death in patients with non-small cell lung cancer. Pharmacoeconomics. 2024;42:109–116. doi:10.1007/s40273-023-01314-2.

11. Palmer S, Borget I, Friede T, et al. A guide to selecting flexible survival models to inform economic evaluations of cancer immunotherapies. Value Health. 2023;26:185–192. doi:10.1016/j.jval.2022.07.009.

12. Latimer NR, Taylor K, Hatswell AJ, et al. An evaluation of an algorithm for the selection of flexible survival models for cancer immunotherapies: pass or fail? Pharmacoeconomics. 2024;42:1395–1412. doi:10.1007/s40273-024-01429-0.

13. Quinn C, Garrison LP, Pownell AK, et al. Current challenges for assessing the long-term clinical benefit of cancer immunotherapy: a multi-stakeholder perspective. J Immunother Cancer. 2020;8:e000648. doi:10.1136/jitc-2020-000648.

