## supplementary_material_file for "Temporal Response and Survival Dynamics After Durvalumab in Stage III NSCLC: A Response-Augmented Continuous-Time Markov Framework"

### Supplementary Methods

#### S1. Estimands and time origin

Time zero is randomization in the durvalumab arm. The primary horizon is 60 months. The four cohort occupancies are ongoing objective response (OR), progression-free control without ongoing objective response (C), progressed disease (P), and death (D). Their restricted mean times are the areas under the corresponding occupancy curves.

The OR state combines complete and partial response. C is a residual progression-free category and is broader than stable disease. All results are cohort averages, not expected durations conditional on entering a state.

#### S2. Survival propagation

For each interval i, the state vector π(t)=[PF(t),P(t),D(t)] is propagated as

$$\pi(t + \Delta) = \pi(t)\ exp(Q_i\Delta)$$

For consecutive PFS landmarks at a and b,

$$\lambda = -log[PFS(b) / PFS(a)] / (b - a)$$

The value of $\mu_i$ is found by bisection so that OS at the end of the interval equals the paired published parameter-derivation landmark. State propagation has a closed-form expression when the transition rates are fixed, but inversion with respect to $\mu_i$ is transcendental. Bisection was therefore used as a stable solution to the resulting monotone one-dimensional calibration problem. Exact reproduction of the six OS/PFS landmarks through month 36 is imposed by construction and is not interpreted as validation. Generator rows sum to zero, off-diagonal rates are nonnegative, and the resulting transition matrices are stochastic.

#### S3. Duration-of-response function

Published conditional probabilities of remaining in response were 81.1%, 70.0%, 58.7%, 53.0%, and 51.1% at response durations of 12, 24, 36, 48, and 60 months. For adjacent landmarks S(a) and S(b), the constant response-exit rate was

$$h = -log[S(b) / S(a)] / (b - a)$$

The median duration of response was not reached. Trapezoidal integration of the five published landmarks gives a descriptive restricted mean duration of response of 40.60 months among responders through 60 months after response onset. This conditional quantity is not the cohort-average response time from randomization.

#### S4. Response-onset distributions

PACIFIC did not publish the individual times of first CR or PR. Four deterministic cumulative detection scenarios were therefore evaluated:

1. **Primary smoothed:** no response before month 2; linear increase to 28.4% at month 12; linear increase to 29.8% at month 24. Linear interpolation is a parsimonious bridge between modeling anchors, not a claim that detection occurs continuously.

2. **Earliest detection:** all eventual responses assigned to month 2.

3. **Assessment scheduled:** equal increments to the early 28.4% ORR at months 2, 4, 6, 8, 10, and 12; equal increments from 28.4% to 29.8% at months 15, 18, 21, and 24.

4. **Full-cohort denominator:** primary timing scaled from 29.8% of 443 measurable-disease patients to 132/476 (27.7%) of the randomized durvalumab cohort.

The primary analysis retains 443 as the denominator because this is the measurable-disease population in which PACIFIC defined and reported ORR. Using all 476 randomized patients implicitly treats those outside the measurable-disease response analysis as nonresponders; it is therefore used as a conservative denominator sensitivity analysis.

For cumulative detected-response distribution $F_r$ and conditional duration survival $S_{DOR}$,

$$R(t) = \int_0^t S_{DOR}(t - u)\, dF_r(u)$$

Numerical convolution used a 0.05-month grid. Integration used the trapezoidal rule on the same grid.

#### S5. Interpretation of response exits

The complement of $S_{DOR}$ is an exit from documented ongoing response, not a uniquely observed transition to another response category. It can reflect loss of CR/PR, subsequent progression, death, or censoring as represented in the published response analysis. Therefore, the convolution partitions marginal PFS occupancy but does not generate a patient-level transition matrix among CR, PR, SD, and PD.

#### S6. Why CR and PR are not allocated separate state times

The initial best-overall-response counts among measurable patients were CR 6 and PR 120. Those counts describe the distribution of best response but provide no CR- or PR-specific onset times, duration distributions, conversions, or relapse times. Applying the 6:120 count ratio to total objective-response time would impose equal duration distributions and create apparently precise state times unsupported by the data. The ratio is therefore retained as descriptive evidence only and is not converted into separate CR and PR time estimates.

#### S7. Partial landmark-uncertainty analysis

A reproducible Monte Carlo analysis generated 3,000 accepted draws using seed 20260729. Each of the six OS/PFS parameter-derivation landmarks was sampled independently on the logit scale using a standard error approximated from its published pointwise 95% interval. Draws were accepted only when OS and PFS were nonincreasing and PFS did not exceed OS at any paired landmark. Initial and mature ORRs were sampled from Jeffreys beta distributions based on 126/443 and 132/443, respectively, with mature ORR constrained not to be lower than initial ORR.

The duration-of-response landmarks and response-onset form were held fixed because their sampling distributions and covariance were unavailable. Consequently, the 2.5th and 97.5th percentiles quantify partial input plausibility under the stated sampling assumptions. They are not formal confidence or credible intervals and probably understate total uncertainty.

#### S8. Numerical verification

The automated test suite verifies:

1. exact reproduction by construction of the six OS and PFS parameter-derivation landmarks;

2. exact reproduction of the five duration-of-response landmarks;

3. nonnegative occupancy in every state;

4. OR occupancy no greater than PFS;

5. sum of the four occupancies equal to one at every grid point; and

6. sum of restricted state times equal to the selected horizon.

### Supplementary Tables

#### Table S1. Aggregate evidence used in the model

| Quantity | Month/duration | Estimate | Denominator | Analytical role |
|---|---|---|---|---|
| OS | 12 | 83.1% | 476 | Parameter derivation |
| OS | 24 | 66.3% | 476 | Parameter derivation |
| OS | 36 | 56.7% | 476 | Parameter derivation |
| OS | 48 | 49.7% | 476 | Temporal validation |
| OS | 60 | 42.9% | 476 | Temporal validation |
| PFS | 12 | 55.7% | 476 | Parameter derivation |
| PFS | 24 | 45.0% | 476 | Parameter derivation |
| PFS | 36 | 39.7% | 476 | Parameter derivation |
| PFS | 48 | 35.0% | 476 | Temporal validation |
| PFS | 60 | 33.1% | 476 | Temporal validation |
| Initial ORR | initial analysis | 28.4% (126/443) | 443 measurable | Response-onset anchor |
| Mature ORR | extended follow-up | 29.8% (132/443) | 443 measurable | Response-onset anchor |
| Remaining in response | 12 after onset | 81.1% | 132 responders | DOR |
| Remaining in response | 24 after onset | 70.0% | 132 responders | DOR |

| Quantity | Month/duration | Estimate | Denominator | Analytical role |
|---|---|---|---|---|
| Remaining in response | 36 after onset | 58.7% | 132 responders | DOR |
| Remaining in response | 48 after onset | 53.0% | 132 responders | DOR |
| Remaining in response | 60 after onset | 51.1% | 132 responders | DOR |

#### Table S2. Piecewise monthly rates

| Component | Interval (months) | Rate per month |
|---|---|---|
| PF→progressed | 0–12 | 0.04876584 |
| PF→progressed | 12–24 | 0.01777647 |
| PF→progressed | ≥24 | 0.01044261 |
| Progressed→death | 0–12 | 0.07859084 |
| Progressed→death | 12–24 | 0.05788036 |
| Progressed→death | ≥24 | 0.04204996 |
| Exit from ongoing response | response duration 0–12 | 0.01745727 |
| Exit from ongoing response | response duration 12–24 | 0.01226564 |
| Exit from ongoing response | response duration 24–36 | 0.01467129 |
| Exit from ongoing response | response duration 36–48 | 0.00851232 |
| Exit from ongoing response | response duration 48–60 | 0.00304228 |

#### Table S3. Primary state occupancy

| Month | Ongoing OR | PF control without OR | Progressed | Death |
|---|---|---|---|---|
| 12 | 26.1% | 29.6% | 27.4% | 16.9% |
| 24 | 22.9% | 22.1% | 21.3% | 33.7% |
| 36 | 19.6% | 20.1% | 17.0% | 43.3% |
| 48 | 16.9% | 18.1% | 13.9% | 51.1% |
| 60 | 15.6% | 15.3% | 11.6% | 57.5% |

#### Table S4. Restricted mean state time

| Scenario | Horizon | Ongoing OR | PF control without OR | Progressed | Death |
|---|---|---|---|---|---|
| Primary | 60 m | 10.92 | 17.69 | 10.71 | 20.69 |
| Earliest detection | 60 m | 11.78 | 16.83 | 10.71 | 20.69 |
| Assessment scheduled | 60 m | 10.91 | 17.70 | 10.71 | 20.69 |
| Full-cohort denominator | 60 m | 10.16 | 18.45 | 10.71 | 20.69 |

All values are months. Rows sum to the stated horizon, subject to rounding.

#### Table S5. Survival temporal validation

| Endpoint | Month | Predicted | Observed | Difference (pp) |
|---|---|---|---|---|
| OS | 48 | 48.9% | 49.7% | −0.76 |
| OS | 60 | 42.5% | 42.9% | −0.38 |
| PFS | 48 | 35.0% | 35.0% | +0.02 |
| PFS | 60 | 30.9% | 33.1% | −2.20 |

#### Table S6. Partial landmark-uncertainty plausibility ranges

| State | Point estimate, months | Median across draws, months | 2.5th–97.5th percentiles, months |
|---|---|---|---|
| Ongoing objective response | 10.92 | 11.12 | 9.87–12.42 |
| PF control without ongoing response | 17.69 | 17.36 | 14.75–20.03 |
| Progressed disease | 10.71 | 10.79 | 8.31–13.66 |
| Death | 20.69 | 20.67 | 18.61–22.55 |

These are partial plausibility ranges, not formal confidence intervals. OS/PFS landmark covariance and duration-of-response uncertainty were unavailable.

#### Table S7. Why six dynamic states are not estimated

| Candidate state or transition | Published support | Treatment in this analysis |
|---|---|---|
| CR and PR prevalence | Counts at the initial analysis | Combined as objective response |
| CR-specific duration | Not reported | Not estimated |

| Candidate state or transition | Published support | Treatment in this analysis |
|---|---|---|
| PR-specific duration | Not reported | Not estimated |
| CR ↔ PR movement | Not reported | Not estimated |
| Pure SD occupancy | Not separable from other nonresponse PF time | Broader residual control state |
| Post-CR relapse timing | Not reported by randomized arm | Not estimated |
| Progressed disease | Identified marginally as OS−PFS | Included |
| Death | Identified as 1−OS | Included |

#### Supplementary Figures

**Figure S1. Response-augmented state occupancy.** Primary proportions over 60 months.

**Figure S2. Sensitivity to response-onset timing and denominator.** Ongoing objective-response occupancy under the primary smoothed, earliest-detection, assessment-scheduled, and full-cohort-denominator scenarios.

**Figure S3. Survival construction and temporal validation.** Model curves, parameter-derivation landmarks through month 36, and validation landmarks at months 48 and 60.

#### Reproducibility package

The `response_state_model` directory contains input CSV files, the executable analysis, automated tests, generated tables, figures, 3,000 uncertainty draws, and their summary. Running `python src/run_model.py` regenerates all outputs with the prespecified seed. Running `python -m unittest discover -s tests -v` executes the numerical checks.

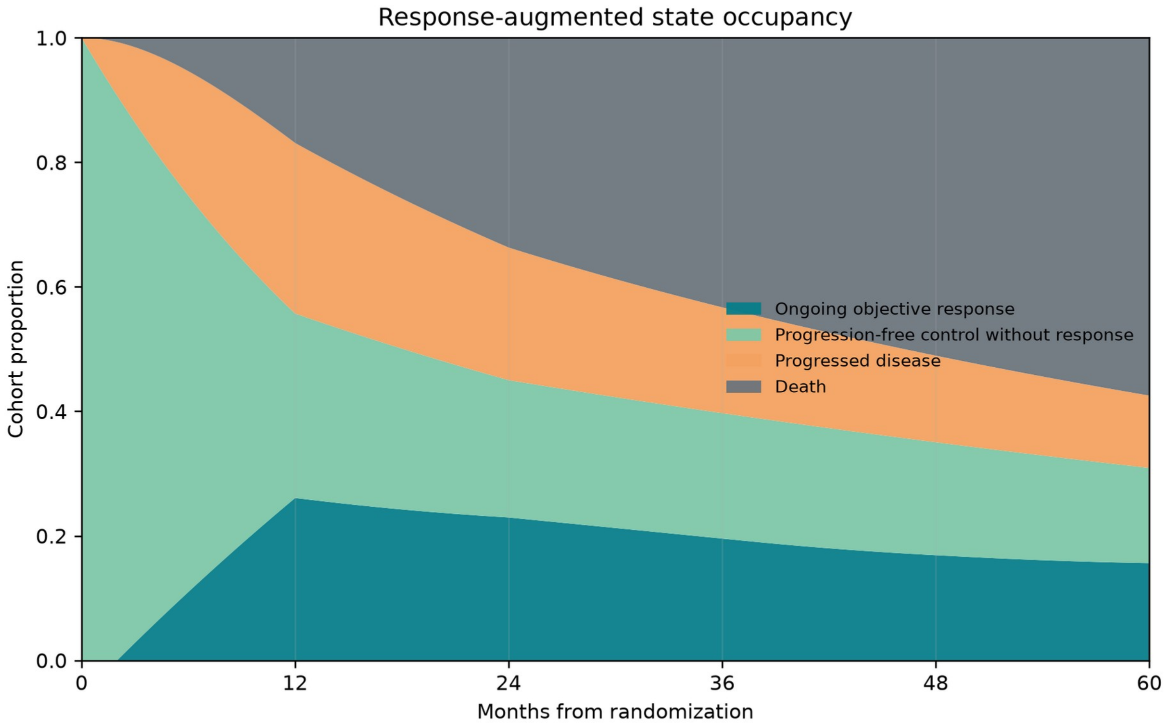


***Figure S1. Response-augmented state occupancy. Primary proportions over 60 months.***

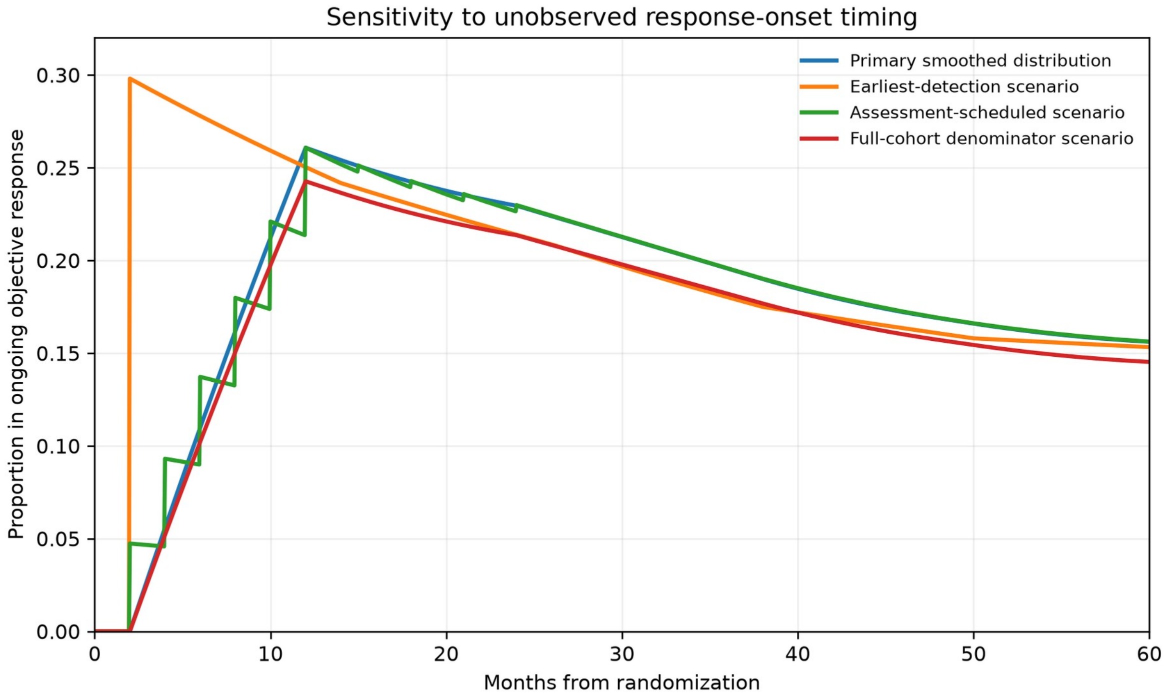


***Figure S2. Sensitivity to response-onset timing and denominator.** Ongoing objective-response occupancy under the primary smoothed, earliest-detection, assessment-scheduled, and full-cohort-denominator scenarios.*

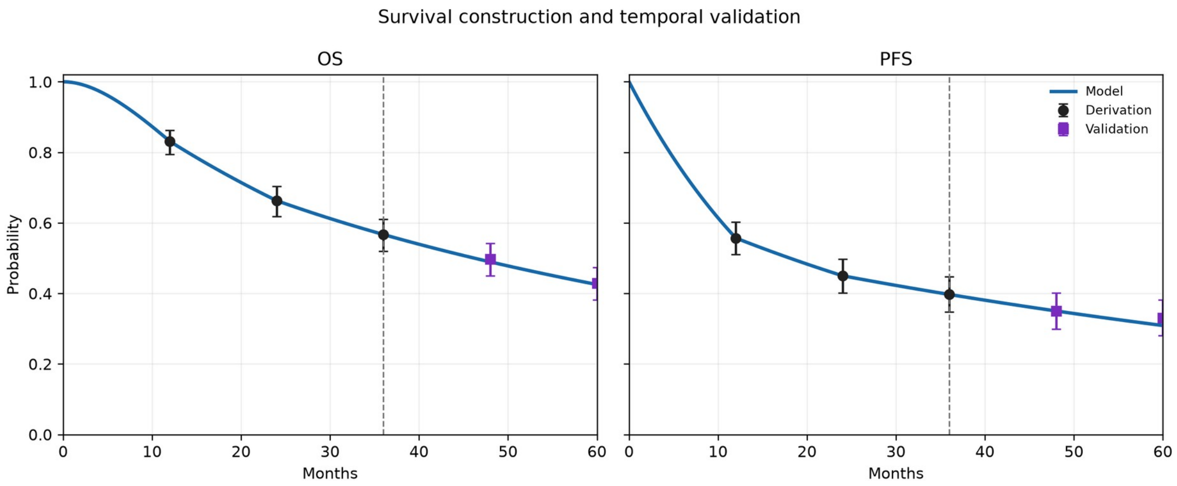


***Figure S3. Survival construction and temporal validation.** Model curves, parameter-derivation landmarks through month 36, and validation landmarks at months 48 and 60.*
